# Racial/ethnic-based spirometry reference equations: Are they accurate for admixed populations?

**DOI:** 10.1101/2021.06.25.21253647

**Authors:** Jonathan Witonsky, Jennifer R. Elhawary, Celeste Eng, José R. Rodríguez-Santana, Luisa N. Borrell, Esteban G. Burchard

## Abstract

**BACKGROUND:** Variation in genetic ancestry among admixed racial/ethnic groups may influence the fit of guideline-recommended spirometry reference equations, which rely on self-identified race/ethnicity.

**RESEARCH QUESTION:** What is the influence of genetic ancestry on the fit of the guideline-recommended racial/ethnic-based spirometry reference equations in populations of genetically admixed children?

**STUDY DESIGN AND METHODS:** Cross-sectional fit of guideline-recommended racial/ethnic-based spirometry reference equations was evaluated in control subjects from case-control studies of asthma. Anthropometry, blood samples, and spirometric measurements were obtained in 599 healthy admixed children, aged 8 to 21-years. Genetic ancestry was estimated using genome-wide genotype data. Equation fit was determined as a mean z-score between -0.5 and 0.5 and assessed in self-identified African American (N = 275) and Puerto Rican (N = 324) children using the distribution to determine cut points of genetic ancestry.

**RESULTS:** For African American children, African American-derived equations fit for predicting FEV_1_ and FVC in those with an African ancestry above the median (81-100%), whereas composite equations for “other/mixed” populations fit for predicting FEV_1_ and FVC in those with an African ancestry below the median (31-81%). Among Puerto Rican children, White-derived equations fit for predicting FEV_1_, and the composite equations fit for predicting FVC for those with African ancestry above the median (21-88%). In contrast, in Puerto Rican children with African ancestry below the median (6-21%), only equations derived in Whites provide an adequate fit.

**INTERPRETATION:** Guideline-recommended spirometry reference equations yielded biased estimates of lung function in admixed populations with high variation of African ancestry. Spirometry is due for reference equations that incorporate genetic ancestry, either for more precise application of the current equations or the derivation and utilization of new equations.

## INTRODUCTION

There has been great debate over the use of racial/ethnic classification in medicine and biomedical research.^1–3^ Presently, pulmonary function testing is one of the few clinical applications where self-reported race/ethnicity is used to define a “normal” range. Normative equations of lung function have been developed by testing large populations categorized by self-reported race/ethnicity.^4,5^ However, many populations have ancestral contributions from multiple continents (i.e., genetically admixed) and, as a result, self-identified racial and ethnic categories may not fully capture an individual’s genetic ancestry.^6–9^ Therefore, using self-reported race/ethnicity may result in misclassifying individuals with respect to the normal range for physiologic lung measures if the latter are more dependent on ancestry.^10^ This error could lead to inaccuracies in evaluating individual pulmonary function.

The two most widely employed sets of spirometry reference equations used in pulmonary function testing, and those recommended by American Thoracic Society and European Respiratory Society (ATS/ERS) guidelines, are the third National Health and Nutrition Examination Survey (NHANES III) equations, derived in White, African American, and Mexican American adults and children aged 8-80 years, and the Global Lung Function Initiative (GLI) equations, derived in White, African American, and North and South East Asian adults and children aged 3-95 years.^4,5,11^ GLI also offers a composite equation—an average of the four population-specific GLI equations—for use in multiracial or unrepresented populations. These guideline-recommended spirometry reference equations adjust for clinically important differences in normal lung function observed between racial/ethnic groups. Selecting an inappropriate reference equation—or ignoring race/ethnicity by using a one-size-fits-all approach—can cause unintended clinical consequences, including errors and delays in disease detection and medical management, misclassification of disease severity, denial of disability claims, and ineligibility for life-saving treatments such as transplants and other surgeries.^12–20^

Several studies have demonstrated that the guideline-recommended lung function equations are well-fitted to populations similar to those in which they were derived.^16,21–25^ However, to date, none of these studies have evaluated equation fit after considering intra-population variation in genetic ancestry, which we have demonstrated is associated with lung function in African Americans and Latinos.^26–28^ Herein, we evaluate the guideline-recommended White, African American, Mexican American, and composite reference equations for two independent genetically admixed populations of African American and Puerto Rican children. Specifically, we assessed guideline-recommended lung function equation fit in each population and investigated their fit after classifying these populations using their genetic ancestry distribution.

## METHODS

### Study Populations

Spirometric predictions and genetic ancestry proportions were calculated using clinical and genetic data from healthy control subjects among two case-control studies of asthma conducted between 2006 and 2014: the Genes-environments & Admixture in Latino Americans (GALA II) study and the Study of African Americans, Asthma, Genes, & Environments (SAGE).^29^ Subjects were aged 8 to 21-years old at the time of recruitment and were healthy with no reported history of lung disease, asthma or allergies, use of medication for allergies, or any symptoms of wheezing or shortness of breath during their lifetime. Parents and grandparents of study subjects self-identified as Puerto Rican in GALA II or African American in SAGE. GALA II subjects were recruited from five urban study centers throughout the U.S. (Chicago, Bronx, Houston, San Francisco Bay Area, and Puerto Rico) and SAGE subjects were recruited from the San Francisco Bay Area.

All participants provided written consent to being in the study. Consent was obtained from all participants 18-years and older and parent/legal guardians of minor participants. The study protocols for both GALA II and SAGE were approved by the University of California, San Francisco (UCSF) Human Research Protection Program Institutional Review Board (IRB) and all institutions participating in recruitment obtained the appropriate approvals from their IRBs for recruitment related activity.

In total, 3,226 children without asthma were enrolled in the two studies (2,538 in GALA II and 688 in SAGE) with complete spirometry and genome-wide genetic data available for 1,049 children. After exclusion criteria (Figure 1) were applied, 599 children remained: 275 African American children and 324 Puerto Rican children.

**Figure 1.**
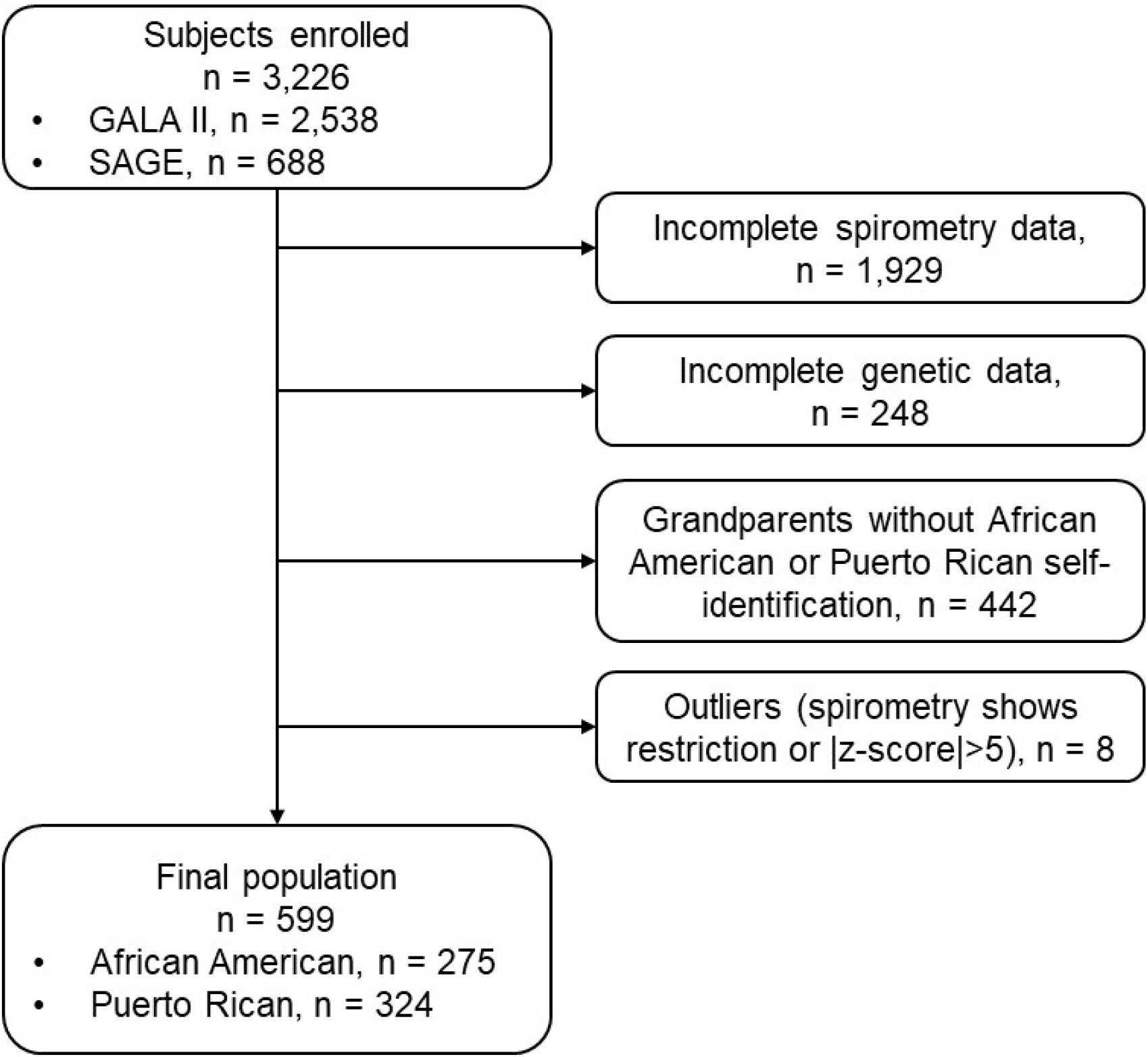
Study population. African American and Puerto Rican children (8-21 years old) recruited as part of parallel case-control studies of asthma from the Genes-environments & Admixture in Latino Americans (GALA II) study and the Study of African Americans, Asthma, Genes, & Environments (SAGE).

### Assessments

Spirometric measurements were collected on all subjects with testing performed in accordance with ATS recommendations.^30^ Samples were genotyped with the Affymetrix Axiom® LAT1 array (World Array 4, Affymetrix, Santa Clara, CA), which includes 817,810 single nucleotide polymorphisms (SNPs). This array was designed to capture genetic variation in African-descent populations such as African Americans and Latinos.^31^ The genetic ancestry of each study participant was determined using an unsupervised analysis in the ADMIXTURE software package, as described elsewhere.^28^

We examined the distribution of genetic ancestry in each population and used the median for African ancestry to classify African American and Puerto Rican populations into two groups, *“greater than median”* and *“less than or equal to median,”* based on African ancestry. Median African ancestry cut points were 81.3% (range: 30.7-100%) for African American children and 21.3% (range: 6.4-87.5%) for Puerto Rican children. Reference equation fit was independently assessed in the African American population (N = 275), the African American population with African ancestry greater than the population’s median (N = 137), the African American population with African ancestry less than or equal to the population median (N = 138), the Puerto Rican population (N = 324), the Puerto Rican population with African ancestry greater than the population’s median (N = 162), and the Puerto Rican population with African ancestry less than or equal to the population median (N = 162).

### Statistical Analyses

Demographic characteristics were compared within each subpopulation using the Student’s t-test for normally distributed continuous variables, the Wilcoxon Rank Sum Test for non-normally distributed continuous variables, and chi-squared statistic for dichotomous variables.

Consistent with previous studies,^32,33^ cross-sectional fit of the equations was determined by calculating FEV_1_ and FVC z-scores. The z-score means for FEV_1_ and FVC were calculated using the NHANES III White, African American, and Mexican American equations and the GLI White, African American, and composite equations. Because all study participants were healthy with assumed normal lung function, an equation was deemed “best fit” when the mean z-score was closest to zero (perfect fit being a mean z-score of zero), and an equation was deemed to have “sufficient fit” if the mean z-score was between -0.5 and 0.5 using two one-sided t-tests (TOST) for equivalence.^21,34,35^ Assessment of significance testing for TOST was performed using 95% confidence intervals and *P*-values.^36^ An equation was deemed appropriate if it provided sufficient fit for both FEV_1_ and FVC. A sensitivity analysis was performed assessing fit using genetic ancestry tertile, quartile, and quintile distributions. To assess for misclassification, the proportion of subjects with observed spirometry data below the lower limit of normal (LLN), which corresponds to the 5^th^ percentile (z-score = -1.65) of predicted values, were also evaluated for FEV_1_ and FVC. Data analyses were conducted using R.^33^

## RESULTS

Table 1 summarizes general characteristics of participants in each racial/ethnic group. Sex, age, height, and weight distributions were comparable across each subpopulation. Lung function, as measured by mean z-scores for FEV_1_ and FVC, was higher for African American (zFEV_1_: 0.41, zFVC: 0.56) and Puerto Rican (zFEV_1_: 0.69, zFVC: 0.55) subpopulations with African ancestry less than or equal to the median than African American (zFEV_1_: 0.29, zFVC: 0.43) and Puerto Rican (zFEV_1_: 0.43, zFVC: 0.23) subpopulations with African ancestry greater than the median. These differences in lung function between subpopulations were statistically significant for the Puerto Rican children but not for the African American children.

**Table 1.**
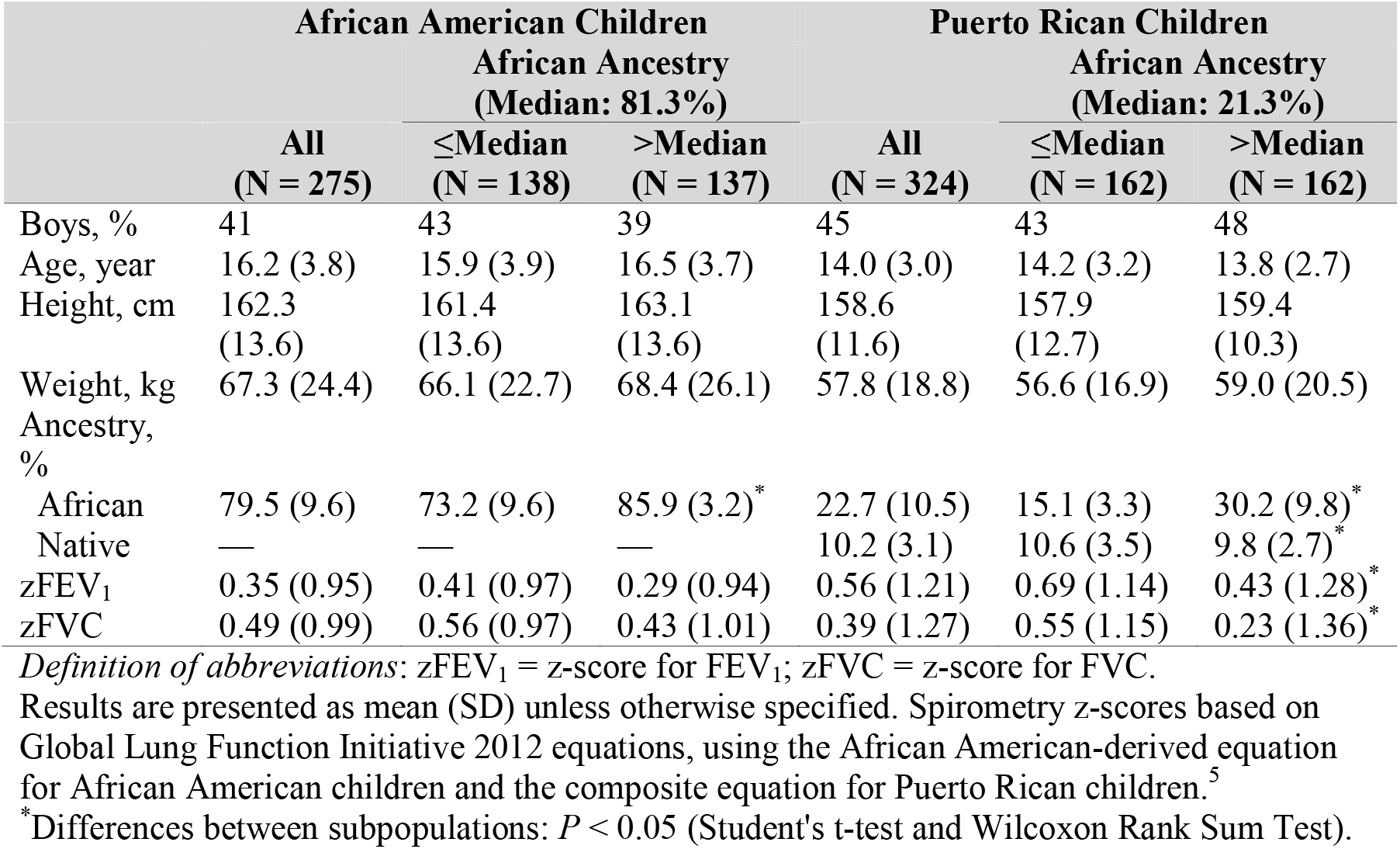
Distribution of selected characteristics for participants in GALA II and SAGE based on race/ethnicity and genetic ancestry categories: 2006-2014.

### Genetic Ancestry Proportions

When comparing subpopulations according to the median distribution, the means (± standard deviation, or SD) of African ancestry were 73.2% (± 9.6%) and 85.9% (± 3.2%) for African American children below and above the median, respectively (Table 1). These values were 15.1% (± 3.3%) and 30.2% (± 9.8%) among Puerto Rican children. Figure 2 shows the European, African, and Native American genetic admixture in the African American and Puerto Rican populations.

**Figure 2.**
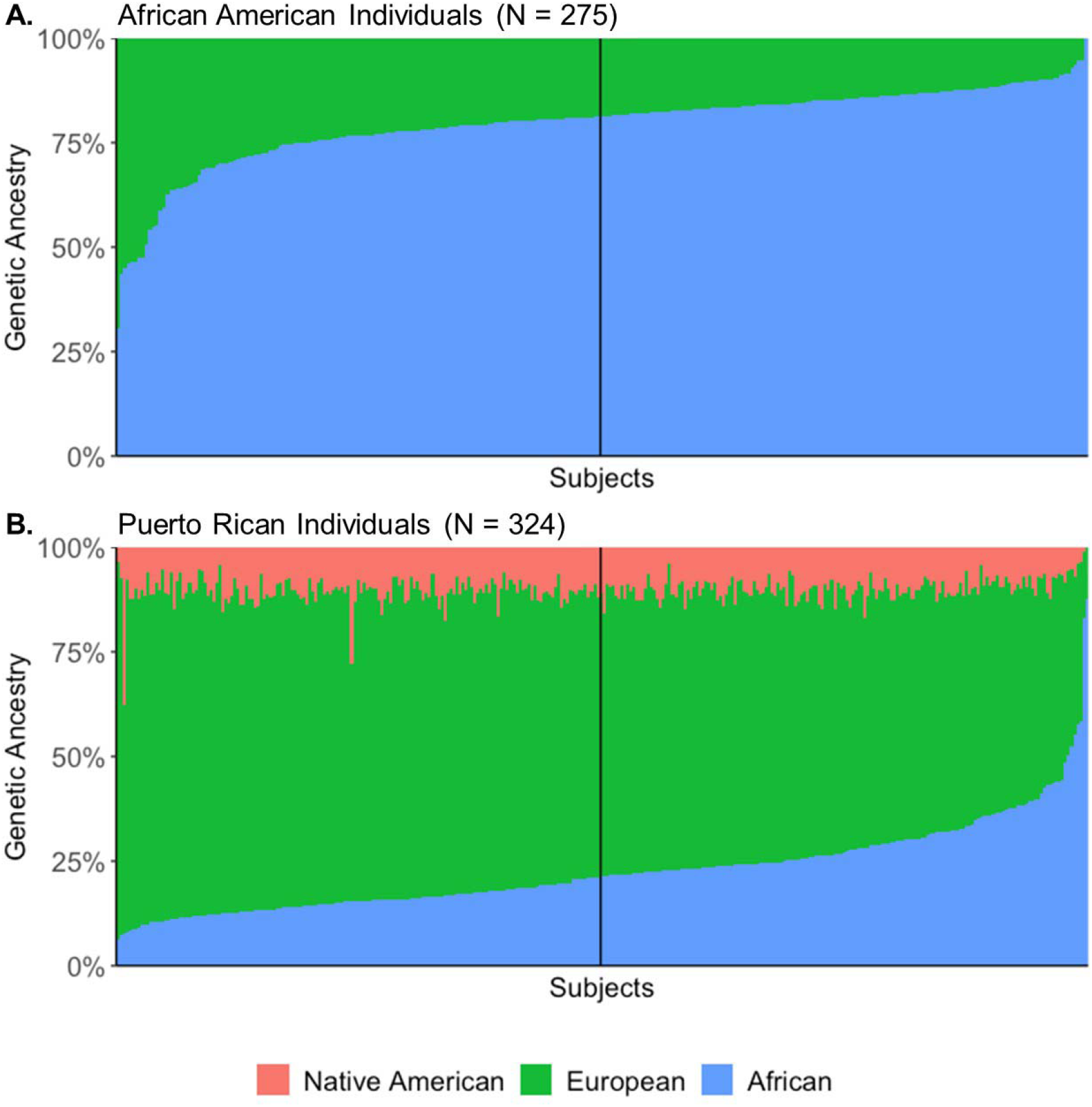
Genetic admixture in the (A) African Americanand (B) Puerto Rican children. The vertical black lines partition each population into paired subpopulations based on proportions of genetic ancestry. The African American and Puerto Rican subpopulations include individuals with African ancestry above and below the median.

### Fit to African American Children

Table 2a provides equation fit to spirometric measurements in the study population and subpopulations of African American children based on genetic ancestry distribution. The NHANES III equations derived in African Americans and the GLI composite equations were appropriate for African American children at the population level, providing sufficient fit for predicting FEV_1_ and FVC. The mean z-scores were 0.17 (95% CI: 0.06 to 0.28) for predicting FEV_1_ (2.5% below the LLN) and 0.34 (95% CI: 0.23 to 0.45) for predicting FVC (1.8% below the LLN) using the NHANES III African American equations, whereas they were -0.32 (95% CI: -0.43 to -0.20) for predicting FEV_1_ (5.8% below the LLN) and -0.14 (95% CI: -0.27 to -0.02) for predicting FVC (6.2% below the LLN) using GLI composite equations. When using the African genetic ancestry median to classify African American children, only the GLI composite equations were appropriate for predicting both FEV_1_ and FVC with mean z-scores of -0.25 (95% CI: -0.42 to -0.09) for FEV_1_ (5.8% below the LLN) and -0.08 (95% CI: -0.25 to 0.10) for FVC (5.8% below the LLN) for African American children classified below the median. In contrast, for African American children with African ancestry above the median, the NHANES III equations derived in African American children were appropriate for predicting FEV_1_ (0.11; 95% CI: -0.04 to 0.26; 2.9% below the LLN) and FVC (0.27; 95% CI: 0.10 to 0.43; 2.9% below the LLN). Figure 3a provides the fit for the NHANES III and GLI equations using other distribution cut points of African ancestry in addition to the median. The figure underscores the poor fit of the equations in African American children when using other African ancestry cut points.

**Table 2a.**
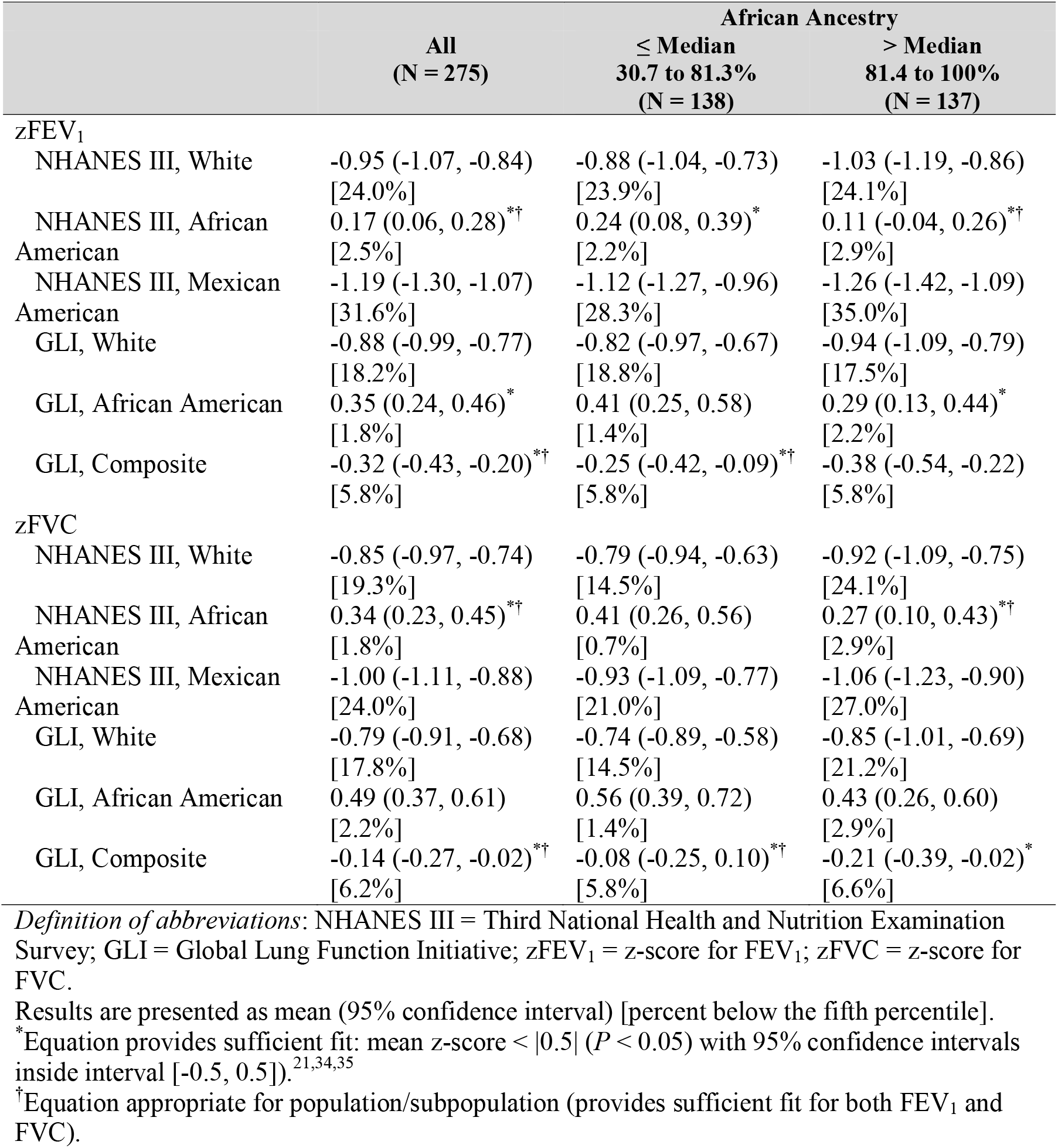
Fit of Spirometry Equation Predictions to Spirometric Measurements in African American Children: 2006-2014

**Figure 3.**
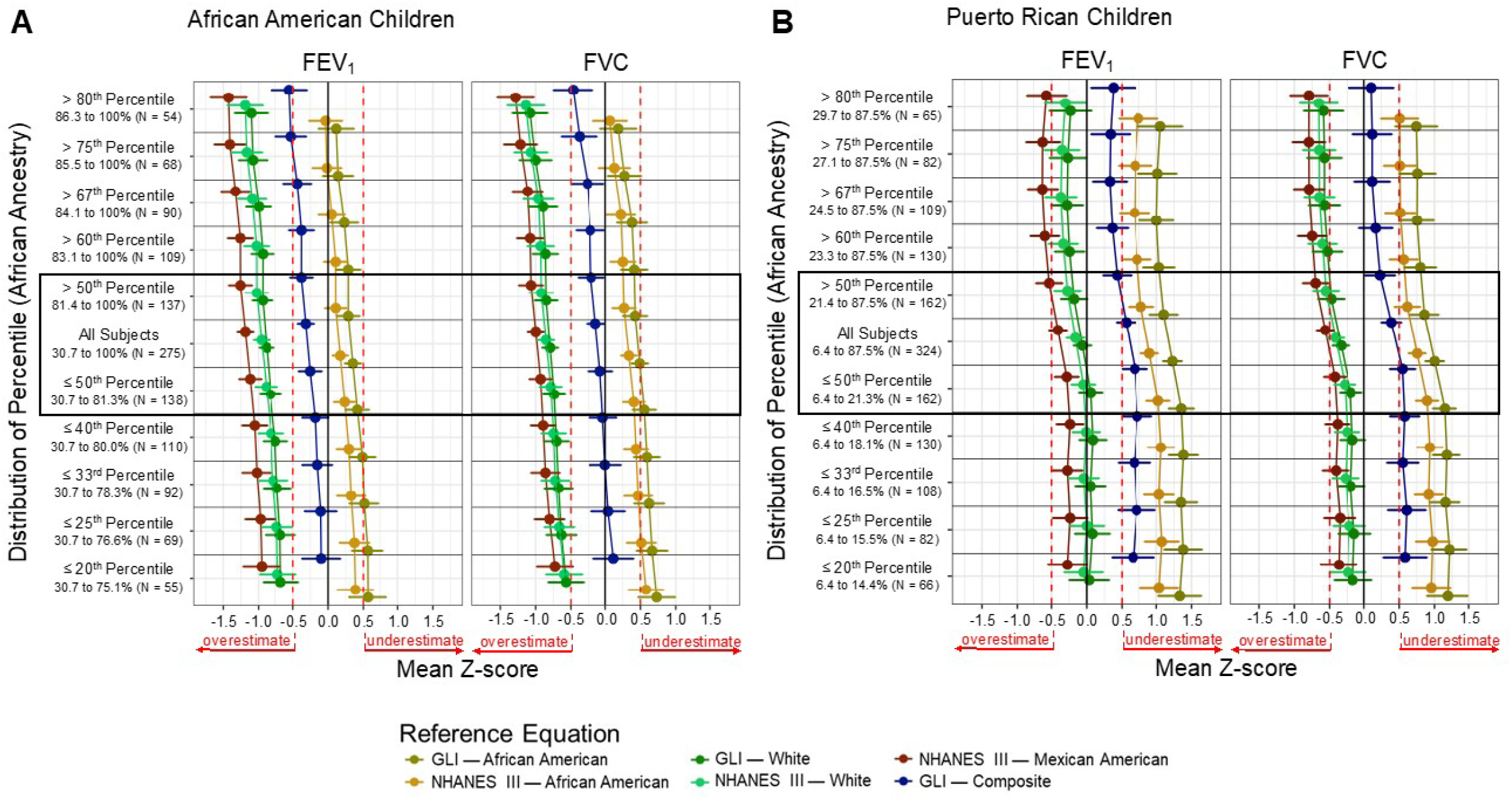
Spirometry reference equation fit for predicting FEV_1_ and FVC in (A) African American and (B) Puerto Rican children with sensitivity analysis showing equation fit in subpopulations with increasingly disparate proportions of genetic ancestry, including ancestry tertiles, quartiles, and quintiles. Dot-and-whisker plots show mean z-score with 95% confidence intervals for each population/subpopulation. *Solid black vertical lines* indicate mean z-score of zero (perfect equation fit). *Dashed red vertical lines* indicate the interval [-0.5, 0.5]) for sufficient equation fit. *Black boxes* outline the primary analysis of the study with sensitivity analysis above and below.

### Fit to Puerto Rican Children

At the population level, the GLI equations derived in Whites were appropriate for Puerto Rican children with mean z-scores of -0.06 (95% CI: -0.19 to 0.06) for predicting FEV_1_ (6.8% below the LLN) and -0.33 (95% CI: -0.46 to -0.21) for predicting FVC (11.1% below the LLN) (Table 2b). For Puerto Rican children with African ancestry below and above the median, the GLI and NHANES III equations derived in Whites fit for predicting FEV_1_. The White-derived equations only fit for predicting FVC among Puerto Rican children with African ancestry below the median. The GLI composite equation fit for predicting FVC in the subpopulation of Puerto Rican children with African ancestry above the median. However, the GLI composite equations underestimated FEV_1_ predictions among those with African ancestry below the median (Figure 3b). In Puerto Rican children with African ancestry below the median, both GLI and NHANES III equations derived in Whites were appropriate for predicting FEV_1_ and FVC.

**Table 2b.** Fit of Spirometry Equation Predictions to Spirometric Measurements in Puerto Rican Children: 2006-2014

|  | All<br>(N = 324) | African Ancestry |  |
| --- | --- | --- | --- |
|  |  | ≤ Median<br>6.4 to 21.3%<br>(N = 162) | > Median<br>21.4 to 87.5%<br>(N = 162) |
| zFEV<sub>1</sub> |  |  |  |
| NHANES III, White | -0.16 (-0.28, -0.04)<sup>*</sup><br>[7.1%] | -0.05 (-0.21, 0.12)<sup>*†</sup><br>[4.9%] | -0.27 (-0.46, -0.09)<sup>*</sup><br>[9.3%] |
| NHANES III, African American | 0.90 (0.78, 1.02)<br>[0.3%] | 1.02 (0.86, 1.18)<br>[0.0%] | 0.78 (0.60, 0.96)<br>[0.6%] |
| NHANES III, Mexican American | -0.41 (-0.53, -0.28)<br>[11.7%] | -0.28 (-0.45, -0.12)<sup>*</sup><br>[9.3%] | -0.53 (-0.72, -0.35)<br>[14.2%] |
| GLI, White | -0.06 (-0.19, 0.06)<sup>*†</sup><br>[6.8%] | 0.06 (-0.11, 0.22)<sup>*†</sup><br>[4.3%] | -0.18 (-0.36, 0.01)<sup>*</sup><br>[9.3%] |
| GLI, African American | 1.23 (1.10, 1.36)<br>[0.3%] | 1.35 (1.18, 1.53)<br>[0.0%] | 1.10 (0.91, 1.30)<br>[0.6%] |
| GLI, Composite | 0.56 (0.43, 0.69)<br>[1.5%] | 0.69 (0.51, 0.86)<br>[0.6%] | 0.43 (0.24, 0.63)<br>[2.5%] |
| zFVC |  |  |  |
| NHANES III, White | -0.41 (-0.53, -0.30)<br>[12.0%] | -0.28 (-0.43, -0.12)<sup>*†</sup><br>[7.4%] | -0.55 (-0.73, -0.37)<br>[16.7%] |
| NHANES III, African American | 0.76 (0.64, 0.87)<br>[0.6%] | 0.89 (0.74, 1.05)<br>[0.0%] | 0.62 (0.44, 0.80)<br>[1.2%] |
| NHANES III, Mexican American | -0.56 (-0.68, -0.44)<br>[15.7%] | -0.42 (-0.58, -0.27)<br>[10.5%] | -0.70 (-0.88, -0.52)<br>[21.0%] |
| GLI, White | -0.33 (-0.46, -0.21)<sup>*†</sup><br>[11.1%] | -0.19 (-0.35, -0.03)<sup>*†</sup><br>[6.8%] | -0.48 (-0.66, -0.29)<br>[15.4%] |
| GLI, African American | 1.00 (0.87, 1.13)<br>[0.6%] | 1.15 (0.98, 1.32)<br>[0.0%] | 0.86 (0.66, 1.06)<br>[1.2%] |
| GLI, Composite | 0.39 (0.25, 0.53)<br>[4.3%] | 0.55 (0.37, 0.73)<br>[1.9%] | 0.23 (0.02, 0.44)<sup>*</sup><br>[6.8%] |

*Definition of abbreviations:* NHANES III = Third National Health and Nutrition Examination Survey; GLI = Global Lung Function Initiative; zFEV<sub>1</sub> = z-score for FEV<sub>1</sub>; zFVC = z-score for FVC.
Results are presented as mean (95% confidence interval) [percent below the fifth percentile].
<sup>\*</sup>Equation provides sufficient fit: mean z-score < |0.5| (*P* < 0.05) with 95% confidence intervals inside interval [-0.5, 0.5]).<sup>21,34,35</sup>
<sup>†</sup>Equation appropriate for population/subpopulation (provides sufficient fit for both FEV<sub>1</sub> and FVC).

## DISCUSSION

Pulmonary function testing is one of the few applications in clinical medicine where self-identified race/ethnicity is used to predict normative values. Our findings confirm previous observations that race/ethnicity-based equations can misestimate lung function after considering intra-population variation in African ancestry.^26^ The influence of genetic ancestry on guideline-recommended equation fit is relevant to African American and Puerto Rican populations in which there are especially wide distributions in the proportions of African ancestry.^37^ Consequently, the guideline-recommended equations may exacerbate inequities in respiratory health, which disproportionately affect the very populations for whom the race/ethnicity-based spirometry equations are the least precise.^38^ As a result, these populations are most prone to underestimated lung function predictions. Misestimated lung function may result in delays of disease detection or overestimated lung function predictions, which can result in unnecessary treatment. This is of critical concern given that African American and Latino/Hispanic children collectively make up approximately 40% of all children in the United States.^39^

Previous studies have validated the use of the GLI equations derived in African American populations for use in African and African American children.^22,40^ Our findings for African American children in this study were consistent with those studies, although only for the total population and ignoring African ancestry variation. After considering the variation in African ancestry, the guideline-recommended equations derived in African American populations were no longer well-fitted for predicting lung function in African American children with African ancestry below the population’s median. Only the GLI composite equation—an equation derived as the average of all GLI data for use in multiracial or unrepresented populations—was appropriately fitted for this subpopulation. This finding suggests that inappropriate spirometry reference equations might be used for as many as half of all African American children, and further illustrates how intra-population variation in African ancestry limits the clinical utility of a single reference equation for an entire genetically admixed group.

In this study, the GLI equations derived in Whites were appropriate for the study population of Puerto Rican children. This was an unexpected finding given that the largest study of reference equation fit for Latino/Hispanic adults found the GLI White equations overestimated lung function among adults who self-identified as Puerto Rican.^18^ The finding was also contrary to the assumption that the GLI composite equation would be most appropriate for Puerto Ricans, a population not represented by the four groups in which the GLI equations were derived. Corroborating prior studies showing that African ancestry is inversely associated with lung function in Puerto Rican individuals,^27,28^ the GLI White equations were not appropriate for Puerto Rican children with African ancestry above the population’s median. In this subpopulation, no equation was appropriately fitted for predicting both FEV_1_ and FVC, suggesting that as many as half of all Puerto Rican children are being misclassified for these measures. The lack of an appropriate equation for Puerto Rican children with more African ancestry is especially problematic considering that asthma prevalence and mortality are highest among Puerto Ricans compared to other racial/ethnic groups.^41,42^ Puerto Ricans have wide-ranging proportions of genetic ancestry and self-identify across the spectrum of the U.S. Census racial categories, making the population ideally suited to the ongoing study of race/ethnicity and genetic ancestry as predictors of lung function.^37,43,44^

There are some important limitations of this study. First, after classifying the populations by genetic ancestry distribution, the sample sizes were below what is recommended for spirometry equation validation studies.^45^ Consequently, equations deemed a poor fit for a subpopulation resulted because this study was underpowered to detect significance when using specified TOST criterion. However, to address this limitation, a sensitivity analysis demonstrated how mean z-scores tended to trend in the serially stratified subpopulations, revealing the continuous influence of ancestry on equation fit. Second, the study did not include the ratio of FEV_1_ to FVC (FEV_1_/FVC), which is the most commonly used outcome to assess airway obstruction. The FEV_1_/FVC ratio was excluded because prior reports suggest that racial/ethnic reductions in FEV_1_ and FVC are proportional and, therefore, the ratio is independent of race/ethnicity.^5,22,46,47^ Third, our findings are not generalizable to adults because the analysis was performed exclusively in children aged 18-21. Lastly, our results were not replicated in other populations of African American and Puerto Rican children, an essential next step in the evaluation of the guideline-recommended equations.

Despite these limitations, this is the first study assessing the influence of genetic ancestry on the applicability of guideline-recommended race/ethnicity-based spirometry reference equations for admixed populations. This study shows that the use of guideline-recommended spirometry reference equations in admixed populations is limited by intra-population variation in genetic ancestry. In addition, this study contributes to the ongoing debate regarding the use of race/ethnicity in clinical algorithms by demonstrating that race/ethnicity, alone, is insufficient for estimating lung function in a large proportion of minority children.^19^ We demonstrate that the use of genetic ancestry data instead of race/ethnicity in lung function equations could improve lung function prediction equations for admixed populations. That said, we recognize that incorporating race/ethnicity captures essential epidemiologic information that relates to lung function and overall clinical outcomes not captured by genetic ancestry.^19^

### INTERPRETATION

This study demonstrates that guideline-recommended spirometry reference equations, which rely on self-identified race/ethnicity, inconsistently fit homogenous racial/ethnic groups after classifying these populations by their genetic ancestry distribution. Recent scientific advances allow for estimates of genetic ancestry to be easily and inexpensively measured as exemplified by direct-to-consumer genetic ancestry testing. Unfortunately, current clinical practice has been slow to adopt and incorporate modern advances from the Human Genome Project, which was started over thirty years ago. Accordingly, spirometry is due for reference equations that incorporate genetic ancestry, either for more precise application of the current equations or the derivation and utilization of new equations. Meanwhile, the increasing availability of genetic data in the clinical setting can be leveraged to guide selection of the most appropriate spirometry reference equation. Our findings suggest that African American children with African ancestry above 80% are best fit by equations derived in African American populations, whereas those with African ancestry below 80% are best fit by the GLI composite equation. Puerto Rican children with African ancestry below 20% are best fit by equations derived in White populations. However, there is no appropriately fitted equation for Puerto Rican children with African ancestry above 20%. As a result, extra caution must be taken when choosing a spirometry reference equation and interpreting its results among racially/ethnically diverse populations. Racially/ethnically diverse individuals might be best served if their spirometric measurements are evaluated using multiple equations, with each result taken in the context of clinical symptoms.

## Data Availability

Biological and phenotypic data analyzed in the current study are available in the dbGAP repository (study accession numbers: SAGE phs00921.v1.p1, GALA II phs001180.v1.p1).

## Author contributions

J. W. contributed to the conception and design of the work, analysis and interpretation of the data, and drafting and revision of the manuscript. C. E. contributed to the acquisition of data. J. R. E., J. R. R.-S., L. N. B., and E. G. B.: contributed to the analysis and interpretation of the data, drafting and revision of the manuscript, and critical review of the manuscript.

## Other contributions

The authors thank the participants and their families for their contribution, as well as the health care professionals and clinics for their support and participation in the Genes-environments & Admixture in Latino Americans Study and the Study of African Americans, Asthma, Genes & Environments. In particular, the authors thank the study coordinator Sandra Salazar and the recruiters who obtained the data: Kelley Meade, MD, Adam Davis, MPH, MA, Lisa Caine, Emerita Brigino-Buenaventura, MD, Duanny Alva, MD, Gaby Ayala-Rodriguez, Ulysses Burley, Elizabeth Castellanos, Jaime Colon, Denise DeJesus, Iliana Flexas, Blanca Lopez, Brenda Lopez, MD, Louis Martos, Vivian Medina, Juana Olivo, Mario Peralta, Esther Pomares, MD, Jihan Quraishi, Johanna Rodriguez, Shahdad Saeedi, Dean Soto, Ana Taveras, and Emmanuel Viera. The authors would also like to thank Andrew Zeiger, Oona Risse-Adams, María G. Contreras, Fernando Picazo, MD, Martin Rofael, MD, Angel C. Y. Mak, PhD, Donglei Hu, PhD, Scott Huntsman, MS, Eric Wohlford, MD, PhD, Sam S. Oh, PhD, MPH, Maria Pino-Yanes, PhD, L. Keoki Williams, MD, MPH, Michael A. Lenoir, MD, Noah A. Zaitlen, PhD, Elad Ziv, MD, Thomas Nuckton, MD, Christopher Gignoux, PhD, and Joshua Galanter, MD for their advice on project conception and analysis.

